# Automated echocardiographic measurements for longitudinal monitoring of ATTR cardiomyopathy: agreement and repeatability analysis

**DOI:** 10.64898/2026.04.07.26349280

**Authors:** Amely Walser, Olivier F. Clerc, Constantin Mork, Andreas J. Flammer, Peder L. Myhre, Rahel Schwotzer, Christoph Gräni, Frank Ruschitzka, Felix C. Tanner, Dominik C. Benz

## Abstract

**Background:** Detection of disease progression is key to personalize treatment strategies in transthyretin cardiomyopathy (ATTR-CM), particularly with emerging therapies. Echocardiography can detect subtle longitudinal changes but is limited by operator dependence. This study evaluates agreement and reproducibility of fully automated, AI-assisted echocardiographic measurements under real-world conditions.

**Methods:** This retrospective study included 62 patients with ATTR-CM undergoing 178 serial annual echocardiograms assessed by a reference cardiologist, a second cardiologist, a novice reader, and a fully automated AI algorithm (Us2.ai). Interrater agreement was assessed using Bland-Altman analysis and intraclass correlation coefficients (ICCs). Intrarater variability for human readers was derived from repeated blinded measurements, with limits of agreement (LoA = mean difference +/− 1.96 x SD) defining the smallest detectable change. AI repeatability was assessed using within-study pairwise differences.

**Results:** AI showed moderate agreement with the reference cardiologist for IVSd and LVEDV (ICC 0.65 and 0.51), with biases of −1.9 mm and −39 mL, respectively. Interrater agreement between cardiologists was good (ICC 0.79 and 0.84) with minimal bias (−0.2 mm and +3 mL). Intrarater variability was moderate to excellent for both cardiologists (LoA 3.0 mm and 43 mL for the reference cardiologist; 2.7 mm and 31 mL for the second cardiologist). AI demonstrated comparable repeatability (LoA 3.6 mm and 37 mL), while the novice showed higher variability (5.1 mm and 61 mL).

**Conclusion:** AI-based measurements demonstrated repeatability comparable to experienced cardiologists. Despite moderate agreement and systematic differences in volumetric assessments, their reproducibility supports automated analysis for longitudinal echocardiographic monitoring.

## Introduction

Transthyretin cardiomyopathy (ATTR-CM) is caused by the accumulation of amyloid fibrils in the myocardium. Disease-modifying therapies, including TTR stabilizers such as tafamidis and acoramidis, as well as TTR silencers such as vutrisiran, significantly reduce morbidity and mortality in most patients with ATTR-CM.^1–3^ However, around 30% of patients still experience disease progression.^1^ With new therapies emerging, understanding the disease course and identifying treatment response early are crucial to address this risk and transform patient management and outcome. Currently, disease progression is monitored through functional capacity, quality of life, or N-terminal pro brain natriuretic peptide (NT-proBNP).^4^ However, these markers change slowly over time and lack specificity for cardiac involvement and disease progression, as they are influenced by various extra-cardiac factors.^4^ Therefore, more precise biomarkers are needed to monitor disease progression, leading to a shift towards cardiac imaging for monitoring disease progression.^5^ Serial echocardiographic assessments, due to their accessibility and cost-effectiveness, are a promising tool for monitoring disease progression, especially when assessed in a core lab.^6^ Even small changes in echocardiographic parameters are shown to have prognostic significance.^7^ However, manual echocardiographic measurements, often by various operators, may lack the precision in daily practice to reliably detect such subtle changes.

Artificial intelligence (AI), with its capacity to minimize measurement variability,^8^ presents a unique opportunity to enhance the precision of serial echocardiographic evaluations. By improving precision, AI could enable detection of subtle longitudinal changes. In this study, we evaluated the potential of automated AI-based echocardiographic measurements for longitudinal assessment in ATTR cardiomyopathy by comparing AI-derived measurements with those obtained by a reference cardiologist, a second cardiologist and a novice reader. Because routine echocardiographic studies typically include multiple acquisitions of the same view, automated analysis may be performed on different images than those used for human measurements, leading to slightly different values across these acquisitions. In this context, variability in AI-derived measurements primarily reflects differences between image acquisitions and selection of image rather than inconsistencies in the measurement algorithm itself. This study therefore deliberately accepted this source of variability to mirror a real-world approach, where repeated measurements are typically performed without systematic documentation of the specific acquisitions used.

We hypothesized that AI-based measurements would show good agreement with experienced human readers and that their repeatability would be comparable to expert measurements while being less variable than the interrater differences observed between human readers. We therefore assessed both agreement between raters and repeatability of measurements to better understand the role of fully automated analysis for real-world longitudinal monitoring.

## Methods

### Participant selection criteria

This longitudinal cohort study, approved by the Zurich Cantonal Ethics Committee (KEK-ZH 2014-0490), included 62 patients with ATTR-CM from the University Hospital Zurich Amyloidosis Registry. Inclusion criteria were a confirmed diagnosis of ATTR-CM and availability of at least two echocardiograms performed 12 months apart. No further exclusion criteria were applied. All participants provided written informed consent. Patients underwent serial echocardiograms between 2013–2024, performed at the University Hospital Zurich or other Swiss hospitals or private practices. Each patient had two echocardiograms 12 months apart (baseline and follow-up), with 25 patients undergoing additional annual follow-up echos, resulting in a total of 178 studies. Agreement analyses were performed on cases with complete data for each respective parameter. New York Heart Association (NYHA) functional class and NT-proBNP were documented within 3 months of the baseline echocardiography.

### Image acquisition and analysis

Comprehensive echocardiograms were acquired according to the recommendations of the American Society of Echocardiography.^9^ Echocardiograms were analysed by four raters: (i) an AI-based algorithm, (ii) a board-certified cardiologist with several years of experience (referred to as “reference cardiologist” hereafter), (iii) a recently board-certified cardiologist (referred to as “cardiologist 2” hereafter) and (iv) a novice medical student without prior echocardiography experience (referred to as “novice” hereafter). Several parameters, including interventricular septal thickness in diastole (IVSd), posterior wall thickness in diastole (PWd), left ventricular (LV) end-diastolic volume (EDV), LV end-systolic volume (ESV), LV ejection fraction (LVEF), and E/e’ ratio were measured. The novice received about 12 hours of training, including initial instruction, self-study, and review of 5 cases with the reference cardiologist. The AI-based algorithm (Us2.ai) algorithm performs fully automated echocardiographic analysis, calculating parameters without manual input.^10, 11^ It uses deep learning to classify video clips into A4C, A3C, or A2C views and filters out low-quality images. A convolutional neural network contours the endocardial border for each frame, automatically detecting end-diastolic and end-systolic frames based on video volume curves, with support from an electrocardiogram when available.^12^

For human raters, LV volumes and ejection fraction (EF) were calculated using the modified Simpson’s rule following the 2015 American Society of Echocardiography (ASE)/European Association of Cardiovascular Imaging (EACVI) recommendations for cardiac chamber quantification.^13^ Echocardiographic measurements by the cardiologists and novice were performed using dedicated software (TOMTEC Image Arena, TOMTEC Imaging Systems GmbH). For AI-human agreement, the single, automatically selected optimal value per parameter from AI were compared to the reference cardiologist. AI within-study repeatability was assessed from multiple measurements automatically generated per parameter across the entire echocardiographic study (i.e. the variability indicates variability in image acquisition rather than AI measurement). To evaluate human intrarater variability, 20 echocardiograms were re-measured by all raters after a 3-month interval. For intrarater variability assessment, original loops were not retained. Instead, the most suitable loop for measurements was selected at baseline and after the 3-month interval separately to ensure conditions comparable to the AI algorithm, which in real-world selects the best echocardiographic loop of each study for measurement. In addition, echocardiographic measurements were compared against cardiac magnetic resonance imaging (CMR). A total of 20 CMR examinations, performed and reported by cardiologists independent of the echocardiographic raters, within a median of 2.5 months (interquartile range (IQR): 0.8, 4.8) of the corresponding echocardiogram, were available. IVSd, PWd, LV EDV and LVEF were compared.

### Statistical Analysis

Continuous variables are summarized as median (IQR), and categorical variables as frequency (percentage).

1. **Interrater agreement** between each rater and the reference cardiologist was assessed using Bland–Altman analysis, reporting the mean bias and 95% limits of agreement (LoA = mean difference ± 1.96×SD of differences). Intraclass correlation coefficients (ICCs) were calculated using a two-way random-effects model for single measures, and relative absolute differences were reported as an additional measure of agreement. ICC values were interpreted according to established thresholds: ICC <0.50 was considered poor, 0.50–0.74 moderate, 0.75–0.89 good, and ≥0.90 excellent agreement.
2. **Agreement with CMR** was assessed for a subset of 20 patients with available CMR performed within a median of 2.5 months of the corresponding echocardiogram, using Bland-Altman analysis and ICCs (two-way random-effects model for single measures).
3. **Intrarater variability** of human raters was assessed by repeated blinded measurements of 20 echocardiograms performed after a 3-month interval by each rater independently, without access to prior measurements. Variability was expressed as limits of agreement (LoA = mean difference ± 1.96×SD of differences), and ICC was calculated using a two-way mixed-effects model for agreement. Based on the principle that LoA encompass 95% of differences between repeated measurements in a stable patient, differences beyond this range are unlikely to reflect measurement variability alone and were therefore defined as the smallest detectable change, and hence considered clinically meaningful.^14^ Within-study repeatability for the AI algorithm was derived from pairwise differences across multiple automated measurements per study as described above, with equivalent LoA calculated from the weighted mean difference and pooled SD.
4. **Clinically meaningful change** between consecutive 12-month echocardiograms was defined as an absolute difference exceeding the rater-specific LoA derived under (2). The McNemar test was used to assess differences in the proportions of clinically meaningful changes between raters, with Bonferroni correction applied for multiple comparisons. For volume parameters, repeatability data were not available from biplane views; therefore 4-chamber views were used consistently across all raters to ensure comparability.
5. **Test-retest repeatability** was assessed using annual changes in echocardiographic parameters over consecutive 12-month intervals as a surrogate measure. To assess whether treatment with TTR-modifying therapy influenced observed changes, intervals were classified as ‘with TTR-modifying treatment’ or ‘without TTR-modifying treatment’ based on treatment status at the interval midpoint, and mean annual changes were compared between groups using Wilcoxon rank-sum tests.

All analyses were performed in RStudio version 4.5.2 using the packages tidyverse, lubridate, stringr, skimr, janitor, irr, DescTools, and rstatix. Two-sided p-values <0.05 were considered statistically significant.

## Results

### Participant Characteristics at Baseline

A total of 62 participants with ATTR-CM (90% male) were included in the study, with a median age of 73 years (IQR: 67–78). ATTRv was present in 7 patients, with Val50Met being the most common variant. Atrial fibrillation was present in 29% of patients, 34% had coronary artery disease and 34% were treated with TTR-modifying treatment (mostly tafamidis) at baseline. (**Table 1**). Echocardiographic parameters by the reference cardiologist at baseline included median IVSd 16 (15-18) mm, LVEDV 124 (110-154) ml and LVEF 49% (40-54) (**Table 2**).

**Table 1:** Patient characteristics at baseline.

| Metric | Value |
| --- | --- |
| **Demographics** |  |
| Age, years | 73 (67-78) |
| Male sex | 56 (90%) |
| Body mass index, kg/m<sup>2</sup> | 26 (24-29) |
| ATTRv | 7 (11%) |
| ATTRwt | 55 (89%) |
| NYHA class | 2 (2-2) |
| **Laboratory Findings** |  |
| NT-proBNP, pg/mL | 1349 (562-2476) |
| **Medication** |  |
| TTR-modifying treatment* at Baseline | 21 (34%) |
| TTR-modifying treatment During Follow-Up | 29 (47%) |
| TTR-modifying treatment Tafamidis | 12 (20%) |
| **Comorbidities** |  |
| Atrial Fibrillation | 18 (29%) |
| CAD | 21 (34%) |
| Carpal Tunnel Syndrome | 12 (19%) |
| Diabetes | 3 (5%) |
| Hypertension | 21 (40%) |
| Neuropathy | 12 (19%) |

*Continuous variables are presented as median (interquartile range) and categorical variables as frequency (percentage). ATTRv, variant transthyretin amyloidosis; ATTRwt, wild-type transthyretin amyloidosis; CAD coronary artery disease; NYHA, New York Heart Association; NT-proBNP, N-terminal pro-B-type natriuretic peptide. Missing data: For all clinical variables except hypertension, missingness was <25%.*
*\*TTR-modifying treatment included TTR stabilizers and silencers.*

**Table 2:** Baseline echocardiographic parameters.

| **Metric** | **Reference**<br>**Cardiologist** | **AI** | **Cardiologist 2** | **Novice** |
| --- | --- | --- | --- | --- |
| IVSd (mm) | 16 (15-18) | 14 (12-16) | 16 (14-18) | 17 (15-19) |
| PWd (mm) | 14 (12-16) | 15 (11-17) | 15 (12-17) | 14 (12-15) |
| LV EDV biplane (ml) | 124 (110-154) | 88 (75-110) | 121 (106-144) | 122 (103-146) |
| LV ESV biplane (ml) | 64 (52-88) | 37 (29-54) | 57 (48-78) | 60 (48-80) |
| LVEF biplane (%) | 49 (40-54) | 56 (50-61) | 51 (44-57) | 50 (44-57) |
| E/e' ratio | 17 (12-21) | 14 (11-17) | 15 (11-19) | 14 (11-18) |

*Variables are presented as median (interquartile range). EDV, end-diastolic volume; ESV, end-systolic volume; EF, ejection fraction; IVSd, interventricular septal thickness at diastole; LV, left ventricular; PWd, posterior wall thickness at diastole.*

### Agreement of AI and Human Raters with the Reference Cardiologist

Interrater agreement metrics are summarized in **Figure 1** (Bland–Altman analysis for IVSd and LVEDV as representative parameters), **Table 3** (interrater ICC values for all parameters), and **Supplemental Figure S1** (relative absolute differences). AI showed the weakest overall agreement with the reference cardiologist. ICC was moderate for IVSd (0.65), LV volumes (0.51 for LVEDV, 0.49 for LVESV), LVEF (0.62), and SV (0.61), and good for PWd (0.81) and E/e′ (0.76). AI showed mean biases of −1.9 mm for IVSd, −39 mL for LVEDV, and +6% for LVEF. Cardiologist 2 showed good agreement with the reference cardiologist across most parameters, with ICCs ranging from 0.70 to 0.88, and minimal biases (−0.2 mm for IVSd, +3 mL for LVEDV, 0% for LVEF). The novice showed good ICC for LV volumes (0.87) and E/e′ (0.82), but only moderate agreement for wall thickness and LVEF, with a systematic overestimation of IVSd (+1.1 mm).

**Figure 1:**
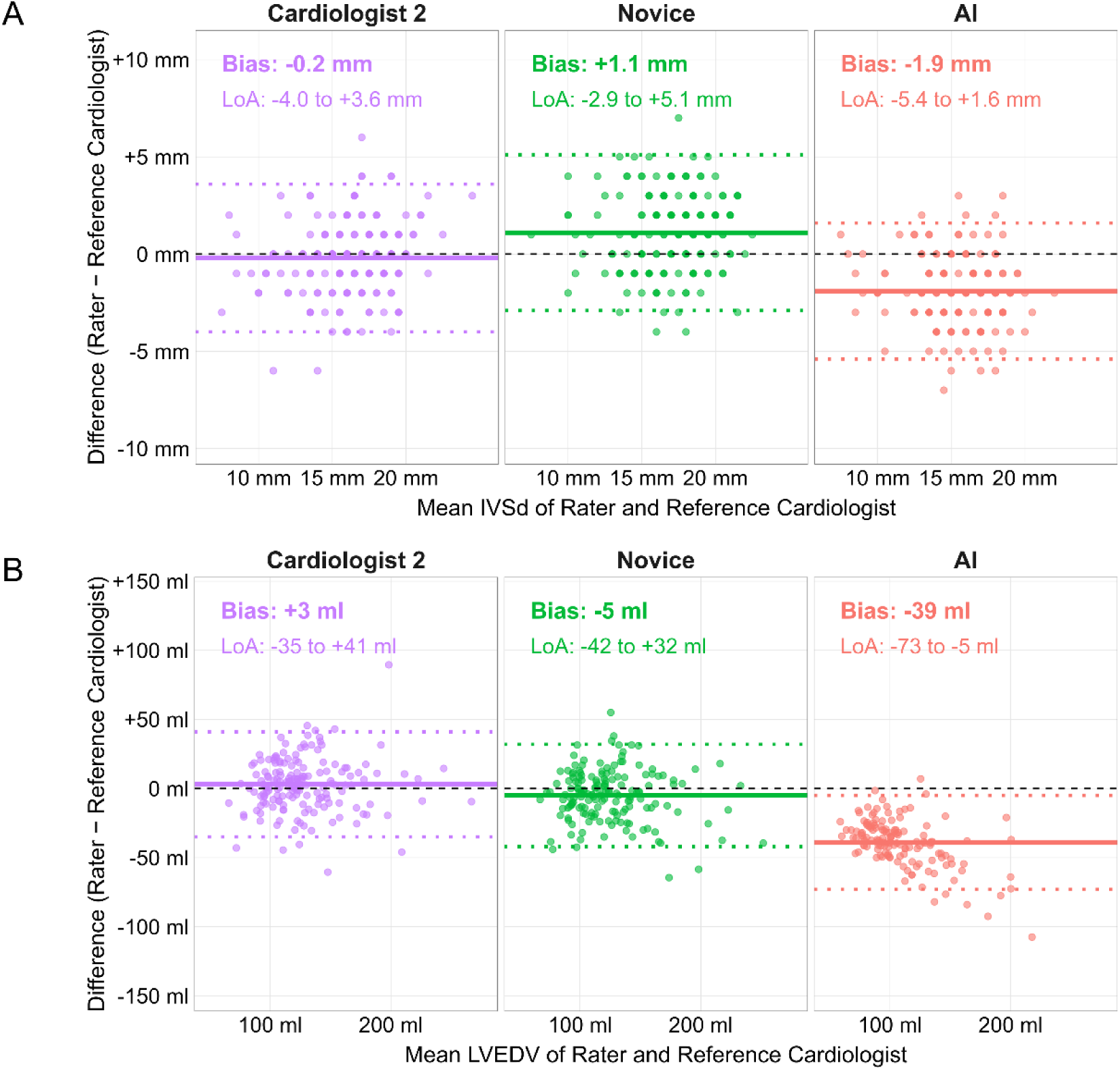
Bland-Altman plots for agreement of AI and human readers with the reference cardiologist. The solid line represents the mean difference (bias), while the dashed lines indicate the 95% limits of agreement (LOA), i.e. ±1.96 standard deviations. Legend entries display bias (lower LOA, upper LOA) for each rater compared to the reference cardiologist. **(A)** IVSd, interventricular septal thickness at diastole; **(B)** LVEDV; left ventricular end-diastolic volume.

**Table 3:** Intraclass correlation coefficients (ICC) for agreement of echocardiographic metrics from AI, cardiologist 2 and novice compared to the reference cardiologist.

| Parameter | AI (ICC 95% CI) | Cardiologist 2 (ICC 95% CI) | Novice (ICC 95% CI) |
| --- | --- | --- | --- |
| IVSd | 0.65 (0.07, 0.85) | 0.79 (0.73, 0.85) | 0.71 (0.48, 0.82) |
| PWd | 0.81 (0.74, 0.86) | 0.71 (0.61, 0.78) | 0.62 (0.51, 0.71) |
| LV EDV biplane | 0.51 (-0.07, 0.82) | 0.84 (0.78, 0.89) | 0.87 (0.81, 0.90) |
| LV ESV biplane | 0.49 (-0.08, 0.81) | 0.83 (0.75, 0.88) | 0.89 (0.82, 0.93) |
| SV biplane | 0.61 (-0.03, 0.84) | 0.75 (0.65, 0.82) | 0.71 (0.60, 0.79) |
| LVEF biplane | 0.62 (0.12, 0.81) | 0.70 (0.61, 0.78) | 0.70 (0.61, 0.78) |
| E/e' ratio | 0.76 (0.38, 0.89) | 0.88 (0.78, 0.93) | 0.82 (0.70, 0.89) |

Red = poor ICC (ICC < 0.50). Orange = moderate ICC (0.50 ≤ ICC < 0.75). green = good ICC (0.75 ≤ ICC < 0.90).
CI, confidence interval; EDV, end-diastolic volume; ESV, end-systolic volume; EF, ejection fraction; ICC, intraclass correlation coefficient; IVSd, interventricular septal thickness at diastole; LV, left ventricular; PWd, posterior wall thickness at diastole; SV, stroke volume.

### Agreement of AI and Human Raters with cardiac magnetic resonance imaging

Agreement metrics between echocardiographic measurements and CMR measurements are summarized in **Figure 2** (Bland–Altman analysis for IVSd and LVEDV), and **Table 4** (ICC values for all parameters). For wall thickness, ICCs indicated poor agreement with CMR for all raters for both IVSd and PWd. All raters slightly underestimated IVSd relative to CMR (biases ranging from −0.4 mm to −3.2 mm), while all raters slightly overestimated PWd (+0.9 mm to +2.3 mm). For LV volumes, ICCs indicated poor agreement with CMR across all raters, with all raters systematically underestimating CMR-derived LVEDV. AI showed the largest bias (−65 ml), followed by the reference cardiologist (−27 ml), and cardiologist 2 (−24 ml). For LVEF, ICCs indicated moderate to good agreement with CMR across all raters. Human raters showed small negative biases (−3% to −5%), consistent with the known tendency of echocardiography to underestimate CMR-derived LVEF, while AI showed minimal positive bias (+1%).

**Figure 2:**
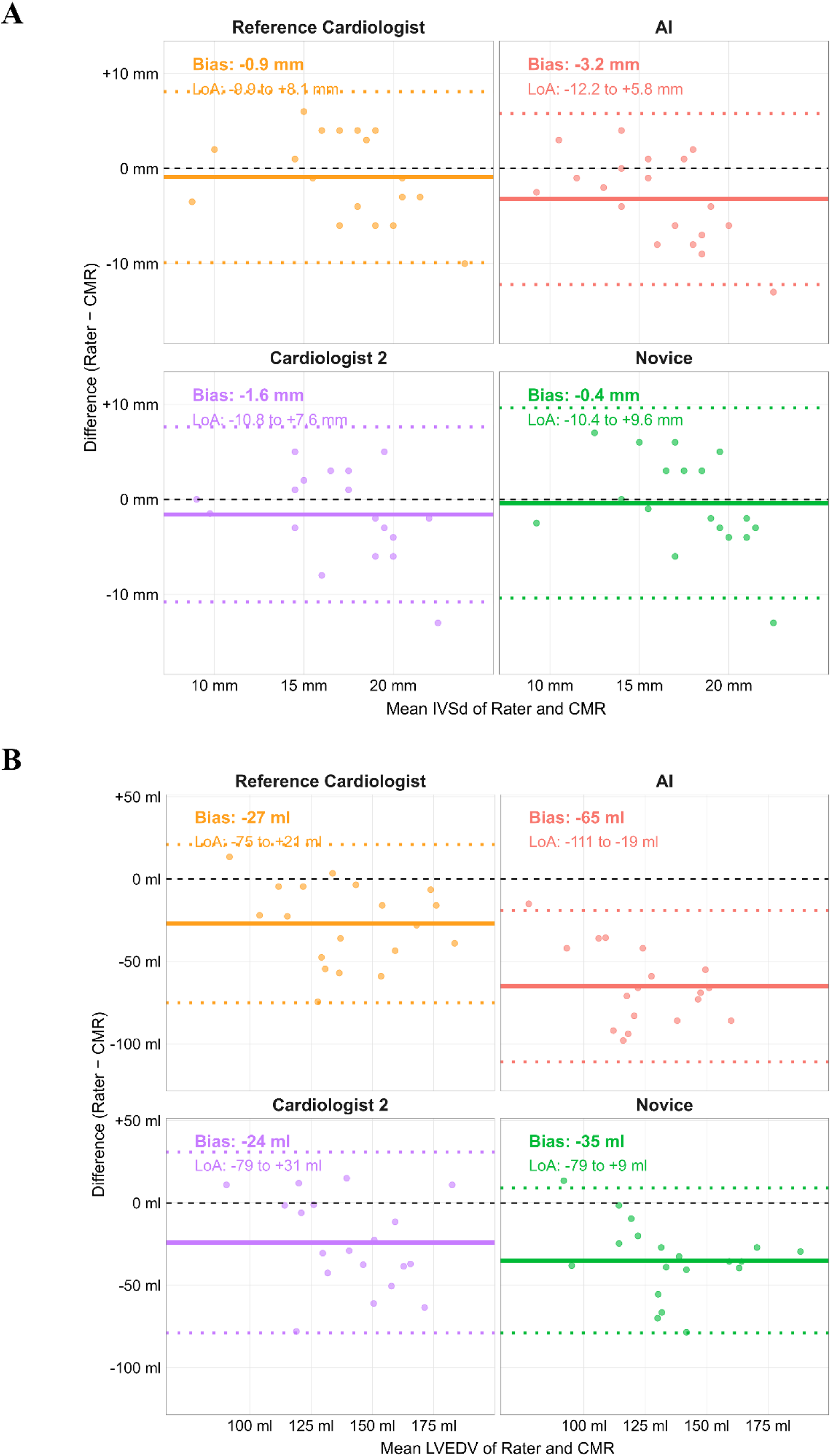
Agreement between echocardiographic measurements and CMR. The solid line represents the mean difference (bias), while the dashed lines indicate the 95% limits of agreement (LOA), i.e. ±1.96 standard deviations. Legend entries display bias (lower LOA, upper LOA) for each rater compared to CMR. **(A)** IVSd, interventricular septal thickness at diastole; **(B)** LVEDV, left ventricular end-diastolic volume.

**Table 4:** Intraclass correlation coefficients (ICC) for agreement of echocardiographic metrics with MR measurements.

| Parameter | Reference Cardiologist<br>(ICC 95% CI) | AI (ICC 95% CI) | Cardiologist 2<br>(ICC 95% CI) | Novice (ICC 95% CI) |
| --- | --- | --- | --- | --- |
| IVSd | 0.46 (0.03, 0.75) | 0.32 (-0.07, 0.65) | 0.39 (-0.03, 0.70) | 0.29 (-0.19, 0.66) |
| LVEDV | 0.43 (-0.09, 0.76) | 0.12 (-0.05, 0.44) | 0.29 (-0.09, 0.63) | 0.37 (-0.11, 0.74) |
| LVEF | 0.77 (0.40, 0.91) | 0.81 (0.57, 0.93) | 0.63 (0.28, 0.83) | 0.63 (0.24, 0.84) |
| PWd | 0.37 (-0.11, 0.74) | 0.39 (-0.07, 0.74) | 0.54 (0.02, 0.83) | 0.42 (-0.09, 0.77) |

Red = poor ICC ( $ICC < 0.50$ ), Orange = moderate ICC ( $0.50 \leq ICC < 0.75$ ), green = good ICC ( $0.75 \leq ICC < 0.90$ ).
CI, confidence interval; EDV, end-diastolic volume; EF, ejection fraction; ICC, intraclass correlation coefficient; IVSd, interventricular septal thickness at diastole; LV, left ventricular; PWd, posterior wall thickness at diastole.

### Intrarater Variability

The repeatability for human measurements was assessed by ICCs (**Table )**. Mean differences and 95% limits of agreement for repeated measurements are summarized in **Supplemental Table S1.** Intrarater variability was moderate to excellent in the reference cardiologist and cardiologist 2, with lower variability than the novice, where intrarater variability was poor for IVSd and moderate for PWd as well as volumes (**Table 5**).

**Table 5:** Intraclass correlations coefficients (ICC) for intrarater variability.

| Parameter | Reference Cardiologist (ICC 95% CI) | Cardiologist 2 (ICC 95% CI) | Novice (ICC 95% CI) |
| --- | --- | --- | --- |
| IVSd | 0.80 (0.58, 0.91) | 0.90 (0.77, 0.95) | 0.47 (-0.03, 0.76) |
| PWd | 0.74 (0.48, 0.88) | 0.80 (0.58, 0.91) | 0.50 (0.07, 0.76) |
| LV EDV biplane | 0.83 (0.55, 0.87) | 0.87 (0.36, 0.96) | 0.71 (0.44, 0.87) |
| LV ESV biplane | 0.80 (0.51, 0.92) | 0.91 (0.75, 0.97) | 0.72 (0.45, 0.87) |
| LVEF biplane | 0.84 (0.66, 0.93) | 0.80 (0.59, 0.91) | 0.86 (0.70, 0.94) |
| E/e' ratio | 0.97 (0.93, 0.99) | 0.95 (0.88, 0.98) | 0.85 (0.67, 0.94) |

Red = poor ICC ( $ICC < 0.50$ ), Orange = moderate ICC ( $0.50 < ICC < 0.75$ ), green = good ICC ( $0.75 < ICC < 0.90$ ), dark green = excellent ICC ( $ICC \geq 0.90$ ).
CI, confidence interval; EDV, end-diastolic volume; EF, ejection fraction; ESV, end-systolic volume; ICC, intraclass correlations coefficients; IVSd, interventricular septal thickness at diastole; LV, left ventricular; PWd, posterior wall thickness at diastole.

Histograms depict differences for repeated measurements by human raters and pairwise differences across multiple automated measurements within the same echocardiographic study by AI (**Figure 3**). For the AI-algorithm, within-study repeatability showed LoA of 3–4 mm for IVSd, 4–5 mm for PWd, and 30–40 mL for LVEDV, which were numerically comparable to those of the expert cardiologists. The novice demonstrated higher variability, with LoA of 5 mm for IVSd, 7 mm for PWd, and 60 mL for LVEDV.

**Figure 3:**
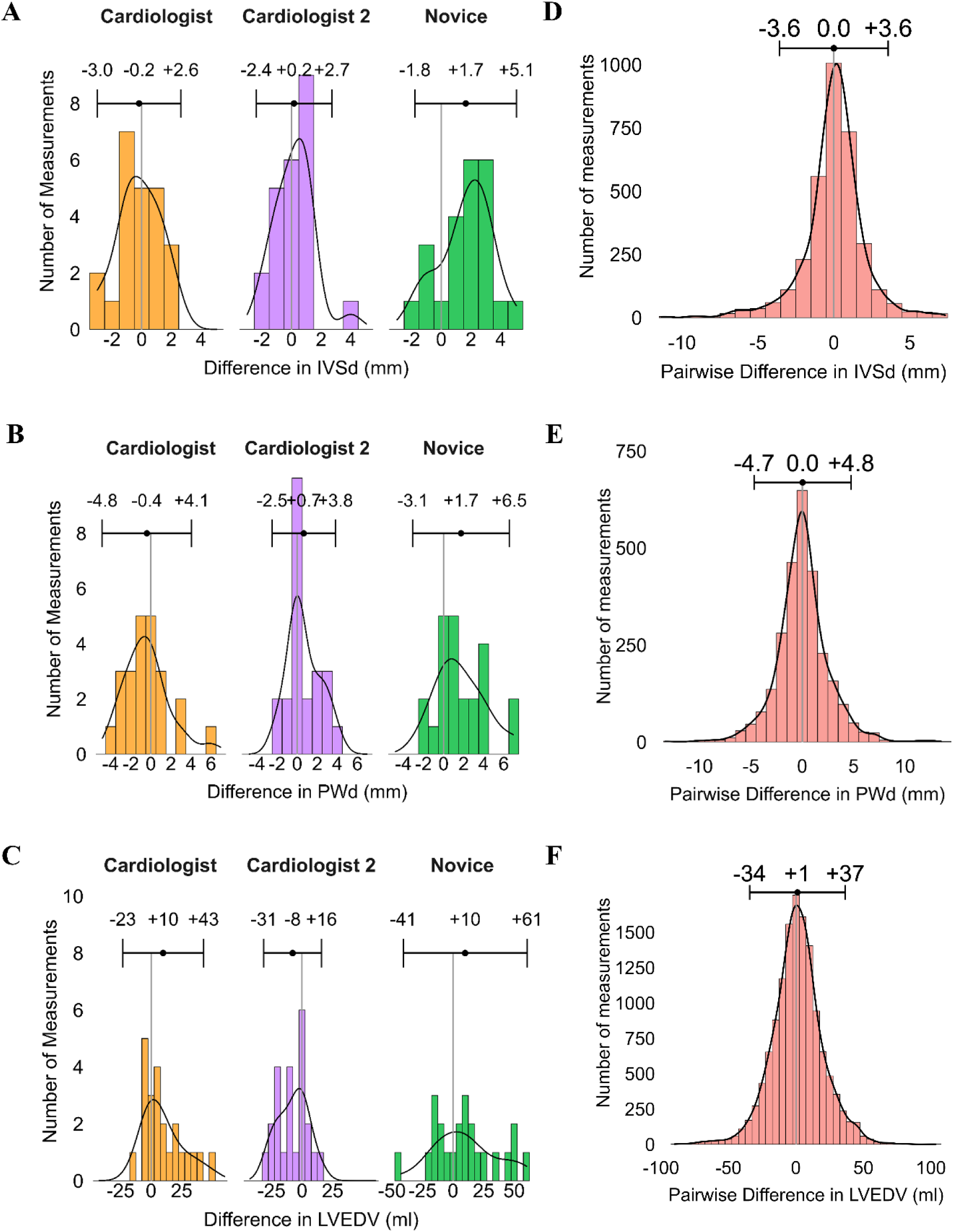
Histogram with differences between two rounds of measurements for human raters (A-C) and with pairwise differences between measurements for AI (D-F). Values shown represent mean ± 1.96×SD of the measurement differences. **(A, D)** IVSd, interventricular septal thickness at diastole; **(B, E)** PWd, posterior wall thickness at diastole; **(C, F)** LVEDV; left ventricular end-diastolic volume.

### Test-Retest Repeatability

Mean annual changes as a surrogate of test-retest repeatability are summarized in **Figure 4** and **Supplemental Table 6**. Overall, temporal changes in the echocardiographic parameters were minimal across all raters, with median changes close to zero for wall thickness parameters and small volume changes for most raters. There were no significant differences in temporal trends for patients treated with vs without TTR-modifying treatment, with consistent findings across all raters (**Supplemental Table 6**).

**Figure 4:**
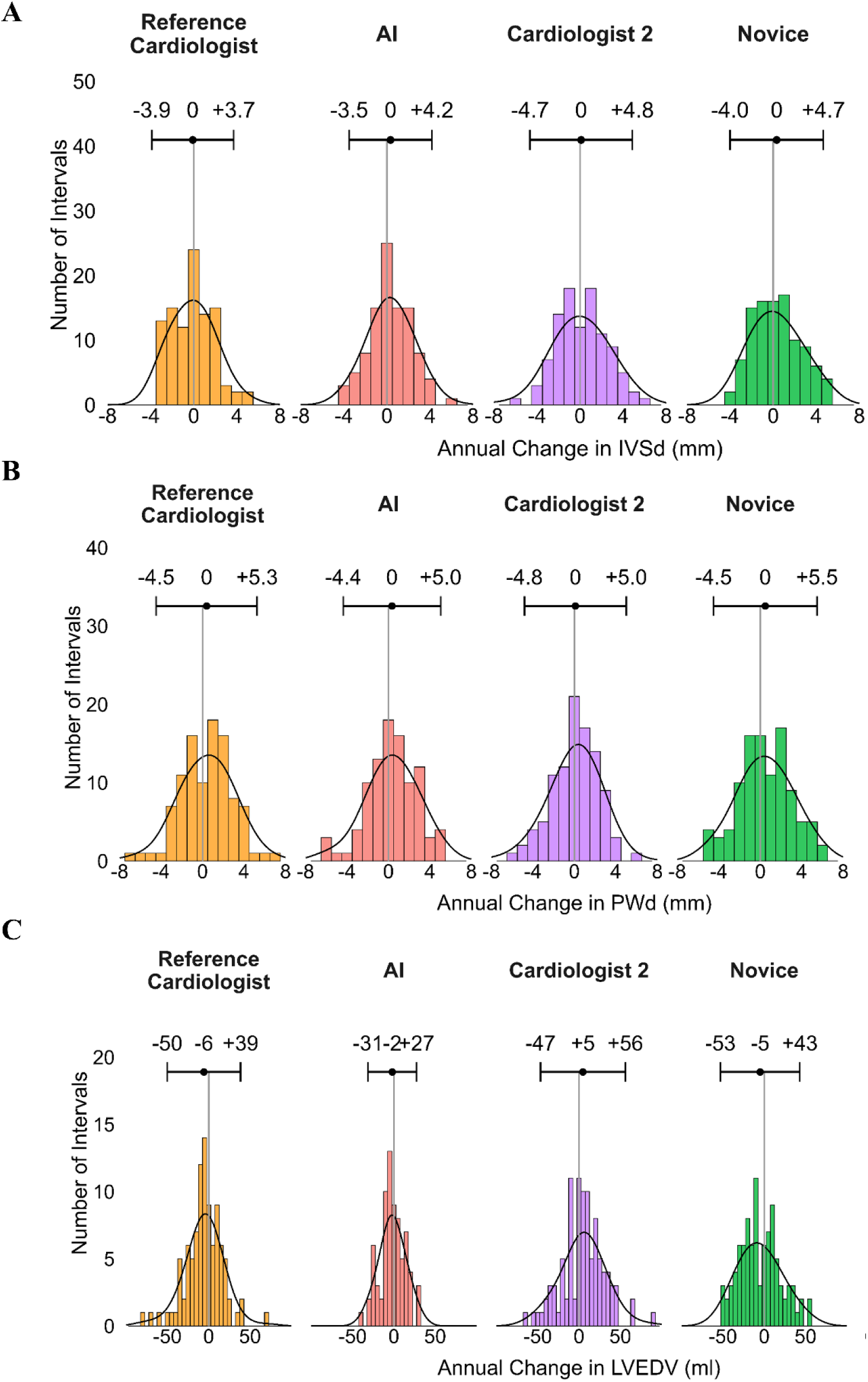
Annual changes. **(A)** IVSd, interventricular septal thickness at diastole; **(B)** PWd, posterior wall thickness at diastole; **(C)** LVEDV, left ventricular end-diastolic volume. Histograms show the distribution of annual changes between consecutive 12-month intervals for each rater as a surrogate of test-retest repeatability. Values above each panel indicate the mean and 95% limits of agreement (±1.96×SD). EDV, end-diastolic volume; ESV, end-systolic volume; IVSd, interventricular septum thickness in diastole; LV, left ventricular; PWd, posterior wall thickness in diastole

Standard deviations of the annual changes were lowest for AI for volume parameters, while they were comparable across all raters for wall thickness parameters. Hence, test-retest repeatability was comparable across all raters, with signs of higher test-retest repeatability of AI for volume parameters (**Figure 4C**).

### Clinically Meaningful Changes

By definition, clinically meaningful changes occurred more often in raters with smaller intrarater variability. The thresholds for clinically meaningful changes as well as the proportion of 12-month intervals reaching clinically meaningful changes are listed and visualized in **Figure 5** and **Supplemental Table S1**. The proportion of clinically meaningful changes did not differ significantly between AI and the reference cardiologist for any parameter (all p>0.05).

**Figure 5:**
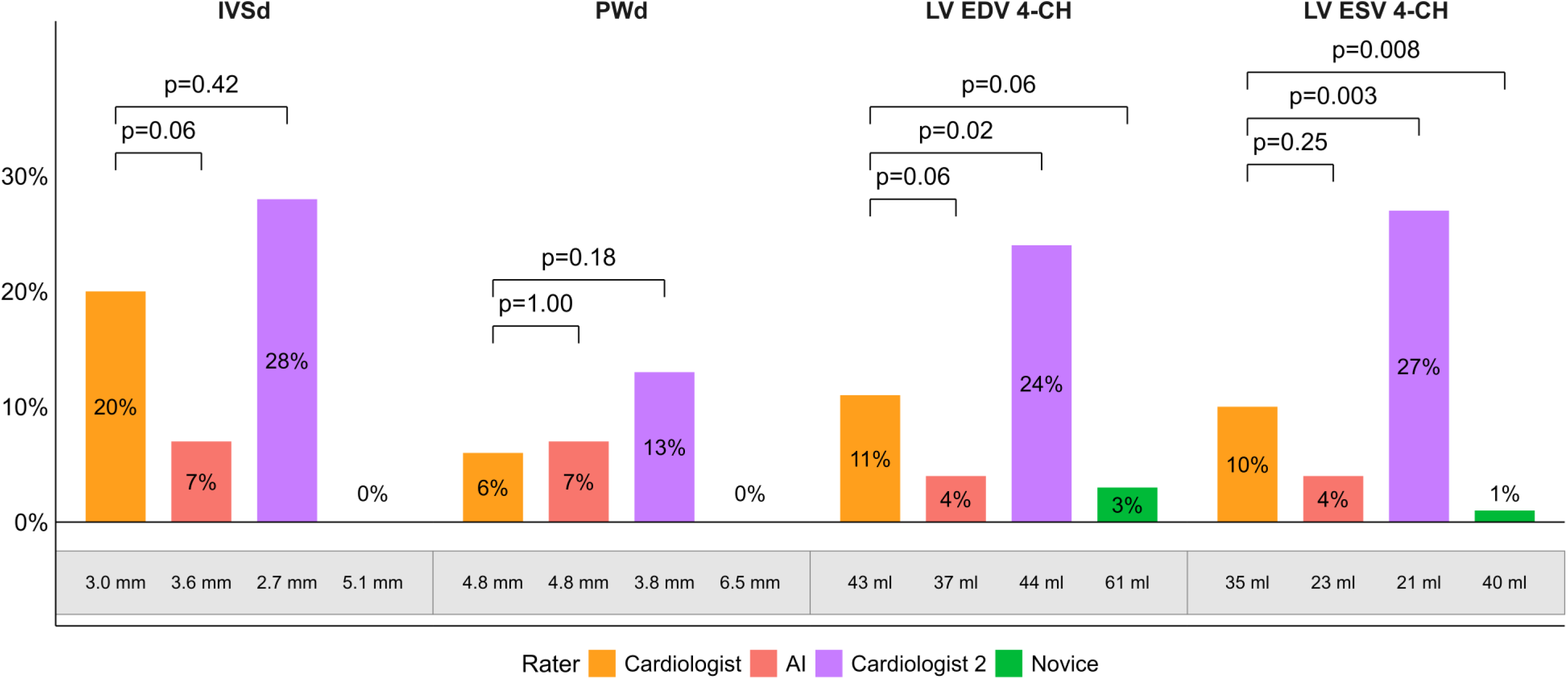
Proportion of 12-months intervals reaching clinically meaningful changes. EDV, end-diastolic volume; EF, ejection fraction; IVSd, interventricular septal thickness at diastole; LV, left ventricular; PWd, posterior wall thickness at diastole. Values in grey box indicate rater-specific thresholds of minimally detectable changes (i.e. 1.96*SD). P-values compare each rater to the reference cardiologist (McNemar’s test, Bonferroni correction for multiple comparisons with p-values >1 capped at 1.00.).

Similarly, cardiologist 2 showed no significant difference for IVSd and PWd (all p>0.05), but detected significantly more changes for LV EDV (p=0.025) and LV ESV (p=0.003). The novice detected no meaningful changes for IVSd or PWd due to high intrarater variability, and significantly fewer changes than the reference cardiologist for LV ESV (p=0.008), with a trend for LV EDV (p=0.063).

## Discussion

Automated echocardiographic measurements using artificial intelligence have the potential to enhance longitudinal assessment of patients with ATTR cardiomyopathy by reducing measurement variability and improving efficiency of serial examinations performed by different operators. In this retrospective analysis, two main findings emerged. First, AI-based measurements demonstrated high repeatability with low variability, comparable to the variability of – and to the agreement between – experienced cardiologists, supporting their potential for reliable longitudinal monitoring of disease progression. Second, moderate agreement between AI and expert measurements and systematic differences in some volumetric measures indicate that human oversight may still be required when applying fully automated measurements in real-world settings with advanced cardiac disease such as ATTR cardiomyopathy. Nevertheless, the consistency of AI-derived measurements suggests that automated analysis may provide a stable framework for tracking changes over time.

### Agreement of automated measurements

AI showed moderate-to-good agreement with the reference cardiologist’s measurements for most parameters, consistent with previous studies on the agreement between AI and human evaluations.^12, 15, 16^ In particular, AI achieved good agreement with the reference cardiologist’s values for PWd, for which the accuracy of the novice was also very good compared to the reference cardiologist. In contrast, for IVSd, both AI and the novice exhibited discrepancies compared with the reference cardiologist’s assessments: while AI consistently underestimated IVSd, the novice tended to overestimate it. This aligns with previous research^16^ and is likely due to challenges in accurately delineating the septal borders from the adjacent right ventricular structures, particularly in ATTR-CM patients with increased septal thickness. Regarding volumetric parameters, both end-diastolic and end-systolic volumes were underestimated by AI, with the underestimation being more pronounced in LVESV. This finding aligns with previous research, which indicated that AI-based measurements exhibited lower reliability and higher bias in LVESV than LVEDV.^12, 17^ This significant underestimation in volumes may be attributed to difficulties in tracing the endocardial border and differentiating between compact myocardium and trabeculations in thicker ventricles.

The agreement we observed was lower than that reported in previous studies involving non-amyloid patients or cohorts with higher LVEF, which generally reported higher AI–human agreement.^10, 15, 17, 18^ Many of these previous studies compared AI-based measurements to human measurements for each individual image or video loop. This contrasts the current study where the AI algorithm performed on multiple images even of the same view. Among studies that did include broader populations, Tromp et al. demonstrated that AI–human differences were similar to or smaller than inter-human variability in a mixed cohort with preserved and reduced LVEF.^10^ The lower agreement in our cohort is likely attributable to three factors specific to our study: first, the real-world setting with unselected patients including advanced disease stages. Second, the particular challenges posed by cardiac amyloidosis, where markedly increased wall thickness complicates automated border delineation. Third, the possibility that the AI-based and human measurements were performed on different images within the same echocardiographic exam.

Several recent studies have examined AI-based echocardiographic tools in cardiac amyloidosis, demonstrating their potential for diagnosis, screening, and disease monitoring. AI-derived measurements have been shown to accurately identify CA in globally diverse cohorts using both multiparametric scoring and deep-learning approaches^19^, and in a real-world unselected population of over 11,000 patients, AI-flagged individuals were 50 times more likely to have confirmed CA.^20^ Furthermore, longitudinal changes in AI-derived LVOT-VTI have been shown to independently predict all-cause mortality in ATTR-CM.^21^ While these studies establish the diagnostic and prognostic potential of AI echocardiography, none specifically examined the agreement and reproducibility of AI-derived measurements against human raters in established ATTR-CM. The only study to specifically examine AI performance in ATTR-CM evaluated fully automated AI-derived LVEF and GLS and found these to be comparable to manual measurements in patients at preclinical stages and at the time of diagnosis.^22^ However, direct comparison with our findings is limited, as that study focused exclusively on LVEF and GLS in patients at earlier disease stages with higher LVEF than ours. The more advanced cardiac involvement in our cohort, with greater wall thickening and more pronounced systolic dysfunction, likely explains the lower AI agreement we observed, consistent with the known reduction in AI accuracy in populations with underlying structural heart disease.^23^ Comparison of echocardiographic measurements with CMR showed that all raters underestimated wall thickness and volumes relative to CMR. For LVEF, where echocardiography is known to consistently underestimate CMR-derived values,^24^ human raters followed this pattern with small negative biases, while AI showed minimal bias, suggesting that despite its underestimation of volumes, AI enables accurate functional assessment.

### Repeatability of measurements

Beyond agreement at a single point in time, high repeatability is critical for long-term monitoring, especially in ATTR-CM, where changes are often subtle and low precision causes missing associations between prognosis and echocardiographic parameters.^25^ Previous work has shown that AI is non-inferior to human intra-observer variability for left ventricular volumes and ejection fraction^18^ and AI models assessing EF by leveraging information across multiple cardiac cycles, were found to be more reproducible than human evaluation.^16, 26^

In our study, the two cardiologists consistently achieved moderate-to-excellent repeatability, while the novice exhibited lower repeatability. AI’s within-study repeatability across various acquisitions of a single study, was comparable to that of the expert cardiologists. Notably, large deviations observed in histograms are attributable to image acquisition variability rather than the measurement algorithm itself. Although variability between acquisitions is common in routine clinical imaging, automated analysis may mitigate the impact of extreme outliers by prioritizing measurements derived from the highest-confidence image acquisitions. Using the mean ± 1.96×SD of repeated measurements as a threshold for clinically meaningful change, AI demonstrated high repeatability across most parameters, outperforming the novice and performing similarly to the cardiologists. These thresholds of clinically meaningful changes were applied for illustrative purposes to demonstrate the clinical implications of differing measurement precision and should not be interpreted as clinically validated criteria for disease progression. Nevertheless, using these illustrative thresholds, the reference cardiologist identified the highest proportion of changes, and AI outperformed the novice.

Apart from one isolated finding for one rater, no differences were detected between intervals with and without treatment. Hence, mean annual changes serve as a surrogate measure of test-retest repeatability: in stable patients (arguably without interval progression or treatment effect within one year), lower standard deviations (SD) in the annual changes indicate higher test-retest repeatability. The SD of annual changes was comparable across all raters for wall thickness parameters. For LV volumes, AI showed the lowest SD of annual changes of all raters, indicating superior test-retest repeatability for automated volume measurements. Taken together, our data demonstrates that AI provides reproducible measurements well suited for serial monitoring, with variability approaching that of a core lab standard with expert cardiologists.

### Clinical Implications

Current guidelines suggest that ≥2 mm increase in LV wall thickness, ≥5% decreases in LV ejection fraction or ≥5 mL decrease in stroke volume are indicative of disease progression.^4^ However, in everyday clinical practice, it is challenging to reliably detect these subtle changes because inter- and intrarater variability limits the precision of echocardiographic assessments. AI offers a promising solution by standardizing measurements and improving consistency in longitudinal evaluations. In this study, the reference cardiologist served as a surrogate core lab, providing standardized assessments which are essential for detecting subtle treatment-related changes. In clinical trials, such core labs standardizing echocardiographic data were critical to assess response to disease-modifying treatment.^6^ Given that AI achieved precision comparable to – and the agreement between – expert cardiologists, it could reduce variability in routine clinical practice and enable more reliable monitoring of treatment responses. In the context of the numerous ongoing therapeutic studies on ATTR amyloidosis, AI-based analysis represents an attractive, cost-effective solution for echocardiographic examinations. In fact, in a randomized controlled trial, integration of AI-based analysis of echocardiography to the clinical workflow enabled sonographers to complete more echocardiograms with less mental fatigue, supporting its role in sustaining high-volume laboratories while maintaining diagnostic quality.^27^

For such fully automated measurements in patients with ATTR-CM, the present study suggests thresholds for minimal detectable changes at 3.6 mm for septal wall thickness, 4.8 mm for posterior wall thickness, 37 mL for LVEDV and 23 mL for LVESV. Nevertheless, in advanced disease cohorts, AI results should be interpreted with caution and supplemented by expert review. It is important that physicians are aware of the limitations of AI-based measurements and critically edit the results, given the potential deviations in measurements to a human expert reader.

### Study limitations

This study has several limitations. First, our assessment of human raters was based on a small number of raters, limiting the generalizability of our findings. Second, within-study repeatability were available only for certain parameters from AI measurements, specifically wall thickness parameters and 4-chamber volume measurements, because the AI software provides multiple repeated measurements for these parameters, whereas LVEF is reported as a single calculated value without individual measurement repetitions. Further, the observed variability, both in human intrarater measurements and AI within-study measurements, may be due to differences in image acquisition and choice of measured image loop rather than the measurement process itself. However, this was deliberately chosen to reflect real clinical practice, where repeated measurements are usually performed without systematic documentation of the specific acquisitions used. Nevertheless, our study also has notable strengths. It expands the comparison of AI-based measurements with human raters to a cardiac amyloidosis cohort at advanced disease stages with a broad range of ejection fractions, thereby highlighting challenges of applying AI in patients with advanced cardiac disease. We also compared echocardiographic measurements against cardiac MR in a subset of patients.

## Conclusion

AI-based echocardiographic measurements demonstrated repeatability comparable to experienced cardiologists, supporting their use for longitudinal monitoring in ATTR cardiomyopathy. While moderate agreement and systematic differences – particularly in volumetric parameters – indicate that human oversight remains important, the reproducibility of AI-derived measurements supports automated analysis as a stable framework for serial evaluation.

## Data Availability

The data referred to in that manuscript is not publicly available.

## Abbreviations

AI: artificial intelligence
ATTR-CM: transthyretin cardiomyopathy
CMR: cardiac magnetic resonance
EDV: end-diastolic volume
ICC: intraclass correlation coefficient
IVSd: interventricular septal thickness in diastole
LoA: limits of agreement
LV: left ventricular
LVEF: left ventricular ejection fraction
PWd: posterior wall thickness in diastole

