## Supplemental Material for "Automated echocardiographic measurements for longitudinal monitoring of ATTR cardiomyopathy: agreement and repeatability analysis"

**Supplement material**

***Supplemental Table S1***: *Repeatability and clinically meaningful changes.*

| | **Mean difference of repeated measurements (± 1.96×SD)** | **Threshold** | **Proportion of clinically meaningful changes** | **p-value (clinically meaningful change vs. cardiologist)[1]** |
|---|---|---|---|---|
| **IVSd** | | | | |
| Reference Cardiologist | -0.2 mm (-3.0, 2.7) | 3.0 mm | 20% | - |
| Cardiologist2 | 0.2 mm (-2.4, 2.7) | 2.7 mm | 28% | 0.417 |
| AI | 0.0 mm (-3.6, 3.6) | 3.6 mm | 7% | 0.056 |
| Novice | 1.7 mm (-1.8, 5.1) | 5.1 mm | 0% | - |
| **PWd** | | | | |
| Reference Cardiologist | -0.4 mm (-4.8, 4.1) | 4.8 mm | 6% | - |
| Cardiologist2 | 0.7 mm (-2.5, 3.8) | 3.8 mm | 13% | 0.179 |
| AI | 0.0 mm (-4.7, 4.8) | 4.8 mm | 7% | 1.00 |
| Novice | 1.7 mm (-3.1, 6.5) | 6.5 mm | 0% | - |
| **LV EDV (4-ch)** | | | | |
| Reference Cardiologist | 10 ml (-23, 43) | 43 ml | 11% | - |
| Cardiologist2 | -8 ml (-31, 16) | 44 ml | 24% | **0.0245** |
| AI | 1 ml (-34, 37) | 37 ml | 4% | 0.0589 |
| Novice | 10 ml (-41, 61) | 61 ml | 3% | 0.0628 |
| **LV ESV (4-ch)** | | | | |
| Reference Cardiologist | 8 ml (-19, 35) | 35 ml | 10% | - |
| Cardiologist2 | -2 ml (-21, 17) | 21 ml | 27% | **0.003** |
| AI | 0 ml (-23, 23) | 23 ml | 4% | 0.25 |
| Novice | 6 ml (-28, 40) | 40 ml | 1% | **0.008** |

[1]*The proportion of clinically meaningful changes was compared between the reference cardiologist and each other rater using McNemar's test with Bonferroni correction for multiple comparisons for paired binary data across all 12-month intervals, p-values >1 were capped at 1.00. In cases where a rater exhibited no clinically meaningful changes (0%), McNemar’s test could not be performed. Values in bold indicate statistical significance.*

*EDV, end-diastolic volume; EF, ejection fraction; ESV, end-systolic volume; IVSd, intraventricular septal thickness in diastole; LAV, left atrial volume; LV, left ventricular; PWd, posterior wall thickness in diastole; 4-ch, 4-chamber view.*

***Supplemental Table S2****: Annual changes as a surrogate of test-retest repeatability.*

| | **Mean Change (overall)** | **Mean Change (without treatment)** | **Mean Change (with treatment)** | **p-value*** |
|---|---|---|---|---|
| **IVSd** | | | | |
| Cardiologist | -0 ± 2 mm | 0 ± 2 mm | 0 ± 2 mm | 0.17 |
| Cardiologist 2 | 0 ± 2 mm | 0 ± 2 mm | 0 ± 3 mm | 0.06 |
| Novice | 0 ± 2 mm | 1 ± 2 mm | 0 ± 2 mm | 0.41 |
| AI | 0 ± 2 mm | 1 ± 2 mm | 0 ± 2 mm | 0.41 |
| **PWd** | | | | |
| Cardiologist | 0 ± 2 mm | 1 ± 2 mm | 0 ± 3 mm | 0.51 |
| Cardiologist 2 | 0 ± 3 mm | 0 ± 2 mm | 0 ± 3 mm | 0.65 |
| Novice | 0 ± 3 mm | 1 ± 2 mm | 0 ± 3 mm | 0.74 |
| AI | 0 ± 2 mm | 1 ± 2 mm | 0 ± 3 mm | 0.08 |
| **LV EDV biplane** | | | | |
| Cardiologist | -6 ± 23 ml | -8 ± 29 ml | -5 ± 19 ml | 0.67 |
| Cardiologist 2 | 5 ± 26 ml | 0 ± 22 ml | 7 ± 28 ml | 0.10 |
| Novice | -5 ± 25 ml | -9 ± 23 ml | -3 ± 25 ml | 0.33 |
| AI | -2 ± 15 ml | -5 ± 16 ml | -1 ± 14 ml | 0.29 |
| **LV ESV biplane** | | | | |
| Cardiologist | -4 ± 16 ml | -7 ± 18 ml | -2 ± 14 ml | 0.17 |
| Cardiologist 2 | 5 ± 21 ml | 2 ± 18 ml | 6 ± 22 ml | 0.29 |

| | | | | |
|---|---|---|---|---|
| Novice | -2 ± 17 ml | -5 ± 18 ml | -0 ± 17 ml | 0.25 |
| AI | -0 ± 9 ml | -2 ± 12 ml | 1 ± 7 ml | 0.83 |

**p-value from Wilcoxon rank-sum test comparing annual changes with vs. without TTR-modifying treatment*

*Values represent mean annual changes ± standard deviation (SD).*

*AI, artificial intelligence; EDV, end-diastolic volume; EF, ejection fraction; ESV, end-systolic volume; LV, left ventricular. IVSd, intraventricular septum end-diastolic.*

***Supplemental Figure S1****: Relative absolute differences compared to the reference cardiologist.*

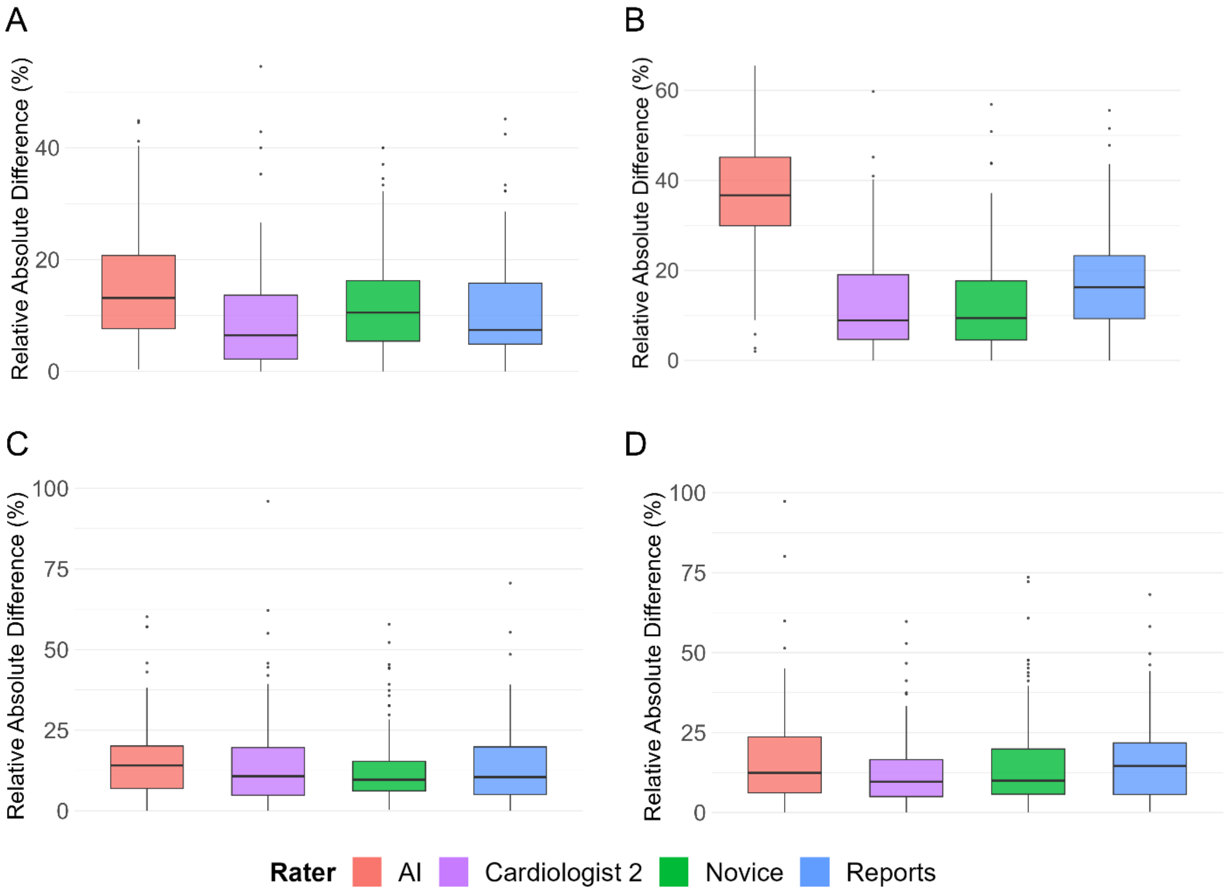


*Relative absolute differences are expressed as percentage of the mean of the two compared raters. **(A)** IVSd, interventricular septal thickness at diastole; **(B)** LVEDV; left ventricular end-diastolic volume; **(C)** LVEF; left ventricular ejection fraction; **(D)** E/e' ratio.*
